# Implementation and validation of a pooling strategy for a sustainable screening campaign for the presence of SARS-CoV-2

**DOI:** 10.1101/2020.08.28.20174946

**Authors:** Daniela Cesselli, Michela Bulfoni, Stefania Marzinotto, Barbara Marcon, Sara Cmet, Anna Rosa Cussigh, Angelica Migotti, Romina Martinella, Corrado Pipan, Antonio Paolo Beltrami, Francesco Curcio

## Abstract

Mass screening aimed at detecting, in asymptomatic subjects, the presence of SARS-CoV-2 is considered a strategic measure for the control of the present pandemic. It allows virus carriers to be identified and quarantined, thus preventing local spread and protecting vulnerable individuals. Although the screening strategy should be determined by the epidemiological situation, the size of the population that can be screened is indeed limited by the availability of resources. Here we present the implementation of an 8-sample pool strategy that relies on protocols, reagents and equipment currently used in clinical diagnostics. The method permitted to identify, with 100% sensitivity, specificity and accuracy, samples with low viral load, being the limit of detection of 11 viral copies extracted from the equivalent of 133μl of nasopharyngeal sample-pool. When the protocol has been applied, as a proof of principle, in a real population of 3592 consecutive nasopharyngeal swabs collected by healthcare providers in asymptomatic subjects, 20 positive pools were detected and in 100% of cases the positive specimens identified. Considering these performances, the 8-sample pool will allow, in populations with an expected positive rate of less than 1%, reducing costs by at least 80%, being a suitable method for a sustainable mass screening strategy in a population of asymptomatic subjects.

## Introduction

The SARS-CoV-2 outbreak represents a challenge for health systems worldwide. As of August 28, according to European Centre for Disease Prevention and Control (ECDC), 24 million people have contracted the virus and 800,000 have died worldwide^1^. In the control of an outbreak, screening, containment and mitigation are considered important strategies^2^. Specifically, screening aims at detecting asymptomatic carriers of the virus in order to guarantee the quarantine of COVID-19 patients to prevent local spread and protect vulnerable individuals ^3^.

Additionally, it informs on the efficacy of the national government measures^3^. However, the screening strategy must be adapted to the national/local epidemiological situation and resources. In fact, although the demonstration of SARS-CoV-2 viral RNA on nasopharyngeal swabs by real-time reverse transcription polymerase chain reaction (RT-PCR) is considered the gold standard method to detect infected subjects, its application in mass screening strongly depends on the availability of diagnosis kits and equipped facilities, especially considering the costs, the worldwide shortage of diagnosis kits and the need to guarantee the execution of diagnostic tests in symptomatic patients^4^.

Pooling is now considered an interesting approach to overcome these problems allowing a sustainable mass screening, even in low income countries^5, 6^. It consists in mixing samples and testing them as a single pool. Subsequently, only the samples of a positive pool are tested singularly. Various pooling strategies have been described and mathematical algorithms to choose the best one are now available^5, 6^. Indeed, to be cost-effective, the pooling strategy must be chosen taking into consideration different factors such as the prevalence of the infection in the tested population, sensitivity and specificity desired and possibility of automation. Pooling has been proven to work for RT-qPCR and, in USA, LabCorp’s COVID-19 molecular test and Quest Diagnostics SARS-CoV-2 RNA have been authorized by the Food and Drug Administration (FDA), under an Emergency Use Authorization (EUA) only, to detect SARS-CoV-2 nucleic acid in pools of, respectively, 5 and 4 upper or lower respiratory specimens^7, 8^.

Here we describe the development and implementation of an 8-sample pooling strategy that allows to detect SARS-CoV-2 mRNA in nasopharyngeal swabs with high sensitivity, specificity and accuracy and reducing costs of reagents by 80% when tested in a population with a prevalence of positive samples up to 1%.

## Materials and Methods

### Sample collection

Ethical approval was obtained from the Medical Research Ethics Committee of the Region Friuli Venezia Giulia, Italy (Consent CEUR-2020-0s- 033). Nasopharyngeal swabs were collected in UTM® tubes (COPAN Diagnostics) by healthcare providers, sent to the Virology laboratory at the University Hospital of Udine and assayed for the presence of SARS-CoV-2 viral RNA by optimized in house protocols^9^.

### Pooling

From each nasopharyngeal sample, 150 μL of medium from 3mL of UTM® tube were taken. Pools were obtained by mixing 8 samples. For determination of sensitivity, limit of detection and specificity, each pool consisted of 7 negative samples, while the 8^th^ sample could be: a negative sample (for specificity assays; n=54), a positive sample (for sensitivity assay; n=170) and a dilution of a positive sample in a negative one (to determine the limit of detection; n=160). In this latter case, 10 different negative pools were added with a positive sample (Cp 15.4) diluted, in the first experiment 10^2^, 10^3^, 5 × 10^3^, 10^4^, 5 × 10^4^, 10^5^, 5 × 10^5^, 10^6^ folds, and, in the second experiment, other 10 different pools were added with the positive sample diluted 10^4^, 5 × 10^4^, 5 × 10^5^, 8 × 10^5^, 10^6^, 2 × 10^6^, 5 × 10^6^, 10^7^ folds. In this way, 160 different extraction were performed. Since each negative pool were composed of 8 different negative samples, we determined whether the different negative-sample composition of the pool could affect the detection of positive samples.

To evaluate the performance in a real population, 3592 consecutive samples, collected by healthcare providers in asymptomatic subjects, were mixed in 8-sample pools (n=449).

### Automated RNA extraction from nasopharyngeal swab

RNA was extracted by the QIAsymphony DSP Virus/Pathogen Kit (Qiagen), using the automated QIAsymphony SP System (Qiagen), following manufacturer’s instructions. Starting from 400 or 800 μL of pool, samples were eluted in 60μL of elution buffer.

### Quantitative Reverse Transcription Polymerase Chain Reaction (RT-qPCR)

SARS-CoV-2 RNA was detected using LightMix® Modular SARS and Wuhan CoV E-gene kit with LightCycler Multiplex RNA Virus Master (Roche) according to the manufacturer’s instructions. Briefly, each reaction mixture contained: 5.4 μL of nucleasefree water, 4 μL of Roche Master, 0.5 μL of reagent mix, 0.1 μL of RT Enzyme, and 10 μL of extracted pool. RT-PCR was performed on a LightCycler 480 II Real-Time PCR System (Roche).

To assay the pooling strategy in a real population of asymptomatic subjects, the 174 pools obtained from 1392 different nasopharyngeal swabs were first analyzed as described above, and, subsequently, only the samples of the positive pools were tested singularly by the DiaSorin Molecular Simplexa™ COVID-19 direct assay system, a real-time RT-PCR system that enables, in 90 minutes, the direct amplification of Coronavirus SARS-CoV-2 RNA from 8 nasopharyngeal swabs (DiaSorin Molecular).

### One-Step Reverse Transcription-Droplet digital Polymerase Chain Reaction (RT-ddPCR)

To determine the absolute number of viral particles in pooled samples, we slightly modified a previously published protocol^9^. Specifically, we used 5’ 6-FAM/3’ BHQ-1^®^-conjugated E-gene1 (Sigma Merck) and One-Step RT-ddPCR Advanced Kit for Probes (Bio-Rad). Briefly, each reaction mixture contained: 5μL of ddPCR™ Supermix for Probes (No dUTP), 900 nM primers and 250 nM probes, 15mM DTT, 20U/μL Reverse Transcriptase, 5 μL sample and nuclease-free water to a total volume of 20 μL. Samples were mixed with Droplet Generator Oil for Probes (Bio-Rad, catalog number 1863005) and droplets were generated on the automated droplet generator QX200™ Droplet Generator (Bio-Rad). PCR amplification was performed by the Veriti^®^ Thermal Cycler (ThermoFisher Scientific). Droplets were read on the QX200™ Droplet Reader (Bio-Rad) and data were analyzed by QuantaSoft™.

#### Limit of Detection (LoD)

LoD has been defined as the lowest analyte concentration at which detection was feasible. LoD was determined by utilizing test replicates of a pool composed of 7 negative samples and different dilution of a sample known to be positive with a Cp=15.4 (see above).

## Results and Discussion

### Sensitivity, specificity and accuracy of RT qRT-PCR detection of SARS-CoV-2 RNA in pool of nasopharyngeal swabs

To establish whether the pooling of samples could affect sensitivity, specificity and LoD of the optimized RT- qRT-PCR assay in use to detect SARS-CoV-2 RNA by LightMix^®^ Modular SARS and Wuhan CoV E-gene kit, a spike-in strategy was used. Specifically, pools of negative samples were first added with a number ranging from n=5 and n=10^4^ of the synthetic positive control supplied with the LightMix^®^ Modular kit. Sensitivity, specificity and accuracy resulted to be 100%, while the limit of detection n=5 copies/reaction, corresponding to a crossing point (Cp) value of 37.19±0.23.

Subsequently, a pool of negative samples was added with the RNA extracted from a patient-derived positive sample diluted from 1 to 10^4^ (corresponding to a number of copies, as evaluated by ddPCR, ranging from 11.0040 to 12), and all dilution were detected by RT qRT-PCR with a Cp value consistent with that obtained using the synthetic positive control.

### Clinical validation

Once assessed, by spike-in samples, the performance of RT qRT-PCR assay in detecting SARS-CoV-2 RNA, we evaluated sensitivity and specificity of the method extracting RNA from 170 positive pools and 54 negative pools. The positive pools were composed by 7 negative samples and 1 positive one, while negative pools were constituted by 8 negative swabs. Although it has been reported that the viral load of symptomatic, pre-symptomatic and asymptomatic patients is similar^22, 23^, 74% of the positive samples used in pools had a Cp value > 30. All negative and positive samples were recognized with 100% sensitivity, specificity and accuracy.

We then evaluated the limit of detection of the method by using an 8-step dilution series of a SARS-Cov-2 positive sample (a nasopharyngeal swab with an assayed Cp 15.4), with 10 replicates each dilution level. Since each replicate was represented by a different pool of 7 negative samples, we excluded that pool composition could affect the efficiency of the detection. The absolute number of copies present in the different pools was assessed by ddPCR. The LoD was equivalent to a dilution corresponding to a Cp value of 37 and an average number of 11 SARS-CoV-2 RNA copies/reaction, corresponding to the RNA extracted by 125μL of pool.

### Use of the pooling in a population of asymptomatic subjects

To assay the pooling strategy in a real population of asymptomatic subjects, 3592 different nasopharyngeal sample were combined in 449 pools that were extracted by QiaSymphony and analyzed by RT qRT-PCR. 20 pools resulted to be positive. The samples constituting the positive pools were analyzed singularly and in all positive pools the positive samples (n=23) were detected by the confirmation tests.

## Discussion

In many countries that have passed the acute phase of the pandemic, the risk of new infections that can cause new SARS-CoV-2 outbreaks is arising. The problem is pressing because, after the lockdown period, it is necessary to guarantee safety in the resumption of production activities and in the reopening of schools. The need to protect risk categories such as healthcare providers, hospitalized patients and the elderly also remains unchanged. Since various studies have highlighted the existence of asymptomatic virus carriers, screening campaigns aimed at identifying and isolating these subjects could substantially contribute to the achievement of these objectives. Defining the prevalence of asymptomatic subjects, the fraction that later develop symptoms, their viral load and their transmission potential are still under investigations^10-15^. However, some mathematical modelling studies hypothesized that asymptomatic subjects can be the major responsible for COVID-19 transmission^16, 17^.

The necessity to screen many subjects can also be part of a surveillance testing aimed at collecting data on incidence and prevalence of the infection to plan and evaluate public health measures^18^.

The effectiveness of mass screening depends on the ability of the test to quickly recognize positive subjects and in the possibility to extend the assay to a large population, according to epidemiological observations^19^. While the first factor relies on the assay sensitivity, the second one is strictly related to the availability of resources, equipment and expertise^19^.

In the case of SARS-CoV-2, pooling testing consists in combining samples to detect SARS-CoV-2 infection: if a pool is negative, then all specimens are assumed negative; if the test is positive or indeterminate, then all the samples in the pool are retested separately^18^. This strategy allows saving reagents and resources, diminishing the time needed to assay a wider population, thus significantly reducing the global costs^18^.

Different pooling strategies have been proposed, based on different pooling schemes and different pool sizes^4-6^. Indeed, to be cost-effective and reliable, the pooling strategy must take into consideration both the virus prevalence in the population and the sensitivity of the assay^20^. Since different phases of a pandemic are characterized by rapidly changing in the prevalence of infected subjects, the testing strategies will need to adapt to these changes in order to be cost-effective, and algorithms have been developed to choose the optimal pool size depending on prevalence rate and test sensitivity^21^. Here we have implemented a pooling strategy based on an 8-sample pool. The pool size chosen, 8 samples, allows, in a population with a virus prevalence in the asymptomatic population of 0.1%, 0.5% and 1% a reduction in the costs of reagents of 87%, 83% and 79%, respectively. Since studies reported that 6-41% of subjects infected with SARS-CoV-2 remain asymptomatic^11-13^, the pool size chosen is suitable for both screening and surveillance testing^18^.

Pooling requires highly sensitive assays in order to detect low positive samples, that, in pool, are further diluted. For this reason, we decided to adopt RT qRT-PCR, a well-recognized method to identify infected patients with high sensitivity and specificity^4, 7 8^. We applied the same procedure (kit, instrument and program) in use in our laboratory for the detection of SARS-CoV-2 E-gene for diagnostic purposes. Moreover, to counteract the "dilution" factor of pooling, we decided to extract RNA by QiaSymphony, an automated procedure that allows not only the purification of RNA but, by choosing the elution volume, its hypothetical concentration up to 13 folds. In this way, loading 10μ1 of pool eluate in each PCR reaction, allowed to analyze the equivalent of RNA extracted from 16 μL of each single nasopharyngeal specimen, thus blunting the dilution factor due to pooling.

Regarding RT qRT-PCR, first we excluded that pooling could affect the PCR-performances. Indeed, by spiking in negative pools extracts a known number of copies of a synthetic positive control, the limit of detection was n=5 copies/reaction corresponding to a Cp value of 37.19±0.23.

Subsequently, we decided to evaluate sensitivity, specificity accuracy and LOD of the optimized assay using pools of nasopharyngeal swabs whose positivity/negativity was first established by gold-standard tests. Both sensitivity, specificity and accuracy resulted to be 100%. Regarding the LOD, it was equivalent to a dilution that corresponded to a Cp value of 37 and an average number of 11 SARS-CoV-2 RNA copies/reaction, as assessed by ddPCR. Since recent reports indicate that the viral load of symptomatic, pre-symptomatic and asymptomatic patients is similar, the performance of the assay could be considered adequate^22, 23^.

Finally, we tested the assay in a real population of 3592 asymptomatic subjects and in the twenty pools resulted positive, we could identify the positive specimens. The prevalence of positivity in the tested population of asymptomatic subjects resulted to be 0.6%, a value that makes 8- sample pooling cost-effective.

In conclusion, the 8-pool strategy implemented allows to detect with high sensitivity and specificity positive samples even when the viral load is low. The procedure relies in instrumentation and expertise already available in the laboratory using RT-qPCR to detect SARS-CoV-2. The possibility to introduce automation both in pool preparation and RNA extraction procedure can further increase the processivity of the system.

Additionally, since the ideal pool can be lower if the prevalence of positivity would increase, it will be easy to reduce the pool size. Indeed, in USA Food and Drug Administration (FDA), under an Emergency Use Authorization (EUA) only, has authorized molecular test to detect SARS-CoV-2 in pools of, respectively, 5 and 4 upper or lower respiratory specimens^7, 8^

In any case, pooling testing procedures are affected by some limitations, that must be taken into account. First, the adequacy of each nasopharyngeal swab included in the pool is not verified before testing. Therefore, inadequate samples will result as negative. Moreover, the dilution in the pool of a specimen with a low viral load, could bring to false negative results. We have implemented a series of corrective measures to reduce this risk, which nevertheless remains.

## Conclusions

We have implemented an-8 sample pool strategy that allows to detect with high sensitivity, specificity and accuracy SARS-CoV-2 infected asymptomatic subjects. This strategy can be already implemented in laboratory using COVID-19 RT-qPCR testing. The platform presents two other major advantages, since it can be easily automated, thus increasing processivity, and it is flexible, since the pool size can easily be lowered if virus prevalence increases. These features make the 8-sample pool combined with COVID-19 RT-qPCR analysis suitable for both screening and surveillance testing.

## Data Availability

All data referred to in the manuscript are available

## Acknowledgements.

We thank technicians, biologists and doctors of the ASUFC molecular biology platform for their support and invaluable help.

